# How Much of the Outcome Improvement after Successful Recanalization is Explained by Follow-up Infarct Volume Reduction?

**DOI:** 10.1101/2023.02.05.23285506

**Authors:** Helge Kniep, Lukas Meyer, Gabriel Broocks, Matthias Bechstein, Friederike Austein, Rosalie McDonough, Caspar Brekenfeld, Fabian Flottmann, Milani Deb-Chatterji, Anna Alegiani, Uta Hanning, Götz Thomalla, Jens Fiehler, Susanne Gellißen, the German Stroke Registry – Endovascular Treatment (GSR – ET)

## Abstract

**Background:** Follow-up infarct volume (FIV) is used as surrogate for treatment efficiency in Mechanical Thrombectomy (MT). In contrast to these assumptions, previous works suggest that MT-related infarct volume reduction has only limited association with outcome comparing MT vs. medical care. It remains unclear to what extent the causal relationship between successful recanalization vs. persistent occlusion and functional outcome is explained by treatment-related reduction in FIV. Results might allow quantification of pathophysiological effects and could improve the understanding of the value of FIV as imaging endpoint in clinical trials.

**Methods:** All patients from our institution enrolled in the German Stroke Registry from 05/2015-12/2019 with anterior circulation stroke, availability of the relevant clinical data and follow-up CT were analyzed. A mediation analysis was conducted to investigate the effect of successful recanalization (Tici≥2b) on good functional outcome (90d mRS≤2) with mediation through final infarct volume.

**Results:** 429 patients were included. 309(72 %) patients had a successful recanalization and 127(39%) achieved good functional outcome. Probability of good outcome was significantly associated with age (OR=0.89,p<0.001), pre-stroke mRS (OR=0.38,p<0.001), FIV (OR=0.98,p<0.001), hypertension (OR=2.08,p<0.05) and successful recanalization (OR=3.57,p<0.01). Using linear regression in the mediator pathway, FIV was significantly associated with ASPECTS (Coefficient(Co)=-26.13,p<0.001), NIHSS admission (Co=3.69,p<0.001), age (Co=-1.18,p<0.05) and successful recanalization (Co=-85.22,p<0.001). Mediation analysis suggest a 23 percentage points (pp) increase of probability of good functional outcome (95%CI:16pp-29pp) in patients with successful recanalization. 56% (95%CI:38%-78%) of the improvement in good outcome was explained FIV reduction.

**Conclusions:** 56% of the improvement of functional outcome after successful recanalization is explained by FIV reduction. Results corroborate established pathophysiological assumptions and confirm the value of infarct volume as imaging endpoint in clinical trials. 44% of the improvement in outcome is not explained by FIV reduction and reflects the remaining mismatch between radiological and clinical outcome measures.

**Trial Registration:** https://clinicaltrials.gov/ct2/show/NCT03356392 (NCT03356392)

## INTRODUCTION

Mechanical thrombectomy (MT) has been shown to improve functional outcome in patients with anterior circulation stroke due to intracranial large vessel occlusion ^1^. It is assumed that the positive treatment effect of MT is caused by fast reperfusion resulting in the salvage and preservation of brain tissue, as indicated by significantly smaller infarct volumes at follow-up imaging compared to patients without successful recanalization ^1,2^. Furthermore, many studies showed a strong association between the extent of ischemic tissue injury assessed at follow-up imaging and functional outcome at 90 days ^3–7^. In consequence, follow-up infarct volume (FIV) has been suggested as an early surrogate of treatment efficacy. In contrast to these assumptions, previous works applying formal testing through mediation analysis suggest that MT-related infarct volume reduction explains only 12% to 14% of the beneficial effect on outcome in patients undergoing MT vs. standard medical care ^2,8–10^. However, these results reflect the explanatory capacity of FIV comparing patients undergoing MT and any recanalization status with patients receiving standard medical care. It remains unclear to what extent the causal relationship between successful recanalization vs. persistent occlusion and functional outcome is explained by treatment-related reduction in FIV. We therefore investigated whether and to what extent FIV mediates the relationship between successful recanalization and functional outcome. We hypothesized that FIV reduction through successful recanalization significantly explains improvement in functional outcome of patients with acute ischemic stroke.

## METHODS

The study was approved by the ethics committee of the chamber of physicians at Ludwig-Maximillians University LMU, Munich (689-15) as the leading ethics committee, in accordance with the Declaration of Helsinki ^11^. Approval by local ethics committees or institutional review boards is obtained for all participating sites according to local regulations.

### Patients

All patients from the University Medical Center Hamburg-Eppendorf (Germany) prospectively enrolled in the German Stroke Registry-Endovascular Treatment (GSR-ET) between June 2015 and December 2019 were analyzed (ClinicalTrials.gov Identifier: NCT03356392). GSR-ET is an ongoing, open label, prospective, multicenter registry of 25 sites in Germany collecting consecutive patients undergoing MT. A detailed description of the GSR-ET study design has been published recently ^12,13^. The main inclusion criteria of GSR-ET are diagnosis of acute ischemic stroke due to large vessel occlusion, initiation of an endovascular procedure for treatment, and age ≥18 years, according to national guidelines. There are no exclusion criteria. For this study, all patients with anterior circulation stroke, availability of relevant clinical data and availability of a follow-up Computed Tomography (CT) of the brain performed 12 hours to 14 days after mechanical thrombectomy (MT) were included. The corresponding author has access to the registry data acquired in the above-mentioned enrolment period. The data that support the findings of this study are available from the GSR-ET registry and the corresponding author, restrictions may apply to the availability of these data.

### Image analysis

Manual segmentation of FIV was performed on follow-up CT scans (slice thickness 4.0 mm) by a senior neuroradiologist with more than 9 years of clinical experience (FA). When multiple follow-up imaging data were available, the latest scan within the time window (12 hours to 2 weeks) was selected for assessment.

### Statistics

Standard descriptive statistics were used for all study end points. Univariable distribution of metric variables was described with mean and interquartile range (IQR), ordinal variables with median and IQR and categorical variables with absolute and relative frequencies. ANOVA tests and Mann-Whitney U tests were used to test for group differences. Categorial variables were compared using chi-squared tests.

Good functional outcome was defined as modified Rankin scale (mRS) 0 to 2 at day 90 after the index procedure. Multivariable logistic regression analysis was performed to identify the independent predictors of good functional outcome. Odds ratios (OR) with 95% confidence intervals (CI) and p-values were calculated for selected variables. Linear regression was used to identify the independent predictors of FIV. Relevant factors were determined using backward variable selection procedures based on the Akaike information criterion (AIC). For the mediation model, all factors significantly associated with good outcome in the logistic regression analysis and all factors significantly associated with FIV in the linear regression analysis were selected, models were additionally controlled for sex and administration of i.v. thrombolysis.

Mediation analysis ^14,15^ was used to evaluate to what extend the reduction of FIV after successful recanalization explains functional outcome at day 90 in patients undergoing MT. Mediation models consider the impact of a mediator variable M that is hypothesized to transmit the influence of independent variables X onto an outcome Y. The underlying regression models were defined according to Mackinnon and Dwyer ^16^. Mediation analysis was conducted in adherence to the requirements defined by Baron and Kenny ^14^ employing algorithms proposed by Imai, Keele and Tingley ^17^ and Imai, Keele and Yamamoto ^18^ that allow estimation of causal mediation effects for linear and nonlinear relationships with continuous and discrete mediators, and various types of outcome variables. Effects were assessed through the average causal mediation effect (ACME) and the average direct effect (ADE) with FIV as mediator variable, good functional status at 90d defined as mRS ≤2 as outcome variable and successful recanalization defined as TICI ≥ 2b as treatment variable. Confidence intervals of the mediation analysis were derived using quasi-Bayesian approximation. 2-sided p-values < 0.05 was considered to be statistically significant. All analyses were carried out using R 3.6.2 with the *mediation* package 4.5.0 ^19^.

## RESULTS

In total, 429 patients fulfilled the inclusion criteria and required data points were available. 309 (72 %) patients received a successful recanalization and 127 (39%) achieved good functional outcome. Patients with good clinical outcome had higher ASPECTS (8 vs. 7, p<0.001), lower FIV (32.3 ml vs 138.9 ml, p<0.001), were younger (67.1 vs. 75.9, p<0.001), had lower mean pre-stroke mRS (3^rd^ quartile 0 vs. 1, p<0.001), lower NIHSS at admission (13 vs. 16, p<0.001), received more often i.v. thrombolysis (72% vs. 52%) and had higher rates of successful recanalization (90% vs. 65%) (Table 1).

**Table 1:** Study cohort clinical characteristics.

| Parameter | Functional Outcome |  | Total (N=429) | p value |
| --- | --- | --- | --- | --- |
|  | good (N=127)<br>90d mRS ≤ 2 | poor (N=302)<br>90d mRS > 2 |  |  |
| Age |  |  |  | < 0.001 |
| - Mean (SD) | 67.1 (12.0) | 75.9 (11.5) | 73.3 (12.3) |  |
| - Q1, Q3 | 60.0, 77.0 | 69.0, 84.0 | 65.0, 82.0 |  |
| Sex (f) | 49 (39%) | 174 (58%) | 223 (52%) | < 0.001 |
| Prestroke mRS |  |  |  | < 0.001 |
| - Median | 0 | 0 | 0 |  |
| - Q1, Q3 | 0, 0 | 0, 1 | 0, 1 |  |
| NIHSS admission |  |  |  | < 0.001 |
| - Mean (SD) | 13 | 16 | 16 |  |
| - Q1, Q3 | 9, 17 | 13, 19 | 12, 19 |  |
| ASPECTS |  |  |  | 0.002 |
| - Median | 8 | 7 | 8 |  |
| - Q1, Q3 | 7, 9 | 6, 9 | 6, 9 |  |
| Hypertension | 81 (64%) | 204 (68%) | 285 (66%) | 0.450 |
| Diabetes | 13 (10%) | 52 (17%) | 65 (15%) | 0.066 |
| Dyslipidemia | 14 (11%) | 40 (13%) | 54 (13%) | 0.527 |
| Atrial fibrillation | 38 (30%) | 111 (37%) | 149 (35%) | 0.175 |
| I.v. thrombolysis | 92 (72%) | 157 (52%) | 249 (58%) | < 0.001 |
| Successful recanalization (Tici ≥2b) | 114 (90%) | 195 (65%) | 309 (72%) | < 0.001 |
| Follow-up infarct volume (ml) |  |  |  | < 0.001 |
| - Mean (SD) | 32.3 (56.7) | 138.9 (161.8) | 107.4 (147.4) |  |
| - Q1, Q3 | 2.6, 30.2 | 18.7, 215.1 | 10.6, 170.2 |  |

Abbreviations: mRS: Modified Rankin Score; SD: Standard Deviation; NIHSS: National Institutes of Health Stroke Scale; ASPECTS: Alberta Stroke Program Early CT Score; Tici: Thrombolysis in Cerebral Infarction.

Results of logistic regression analysis suggest that probability of good outcome was significantly associated with age (OR = 0.89, p<0.001), pre-stroke mRS (OR = 0.38, p<0.001), FIV (OR = 0.98, p<0.001), hypertension (OR = 2.08, p<0.05) and successful recanalization (OR = 3.57, p<0.01). ASPECTS (OR = 1.00), NIHSS at admission (OR = 0.96) and i.v. thrombolysis (OR = 1.79) did not reach significance at p-value < 0.05 (Figure 2). Using linear regression in the mediator pathway, FIV was significantly associated with ASPECTS (Coefficient (Co) = −26.13, p<0.001), NIHSS at admission (Co = 3.69, p<0.001), age (Co = −1.18, p<0.05) and successful recanalization (Co = −85.22, p<0.001). According to these results, the loss of one ASPECTS point led to a mean increase of FIV by 26 ml and successful recanalization was associated with a lowering of FIV by 85 ml (Figure 1).

**Figure 1:**
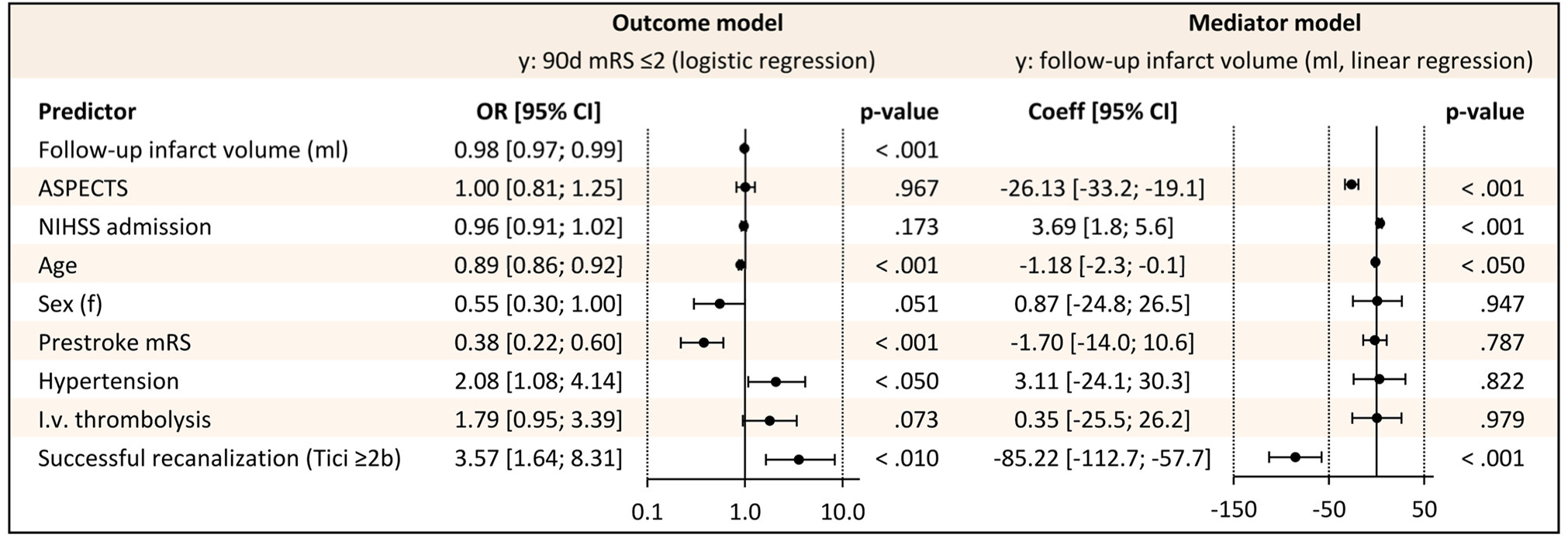
Mediation model regression analysis. Abbreviations: mRS: Modified Rankin Score; SD: Standard Deviation; NIHSS: National Institutes of Health Stroke Scale; ASPECTS: Alberta Stroke Program Early CT Score; Tici: Thrombolysis in Cerebral Infarction.

**Figure 2:**
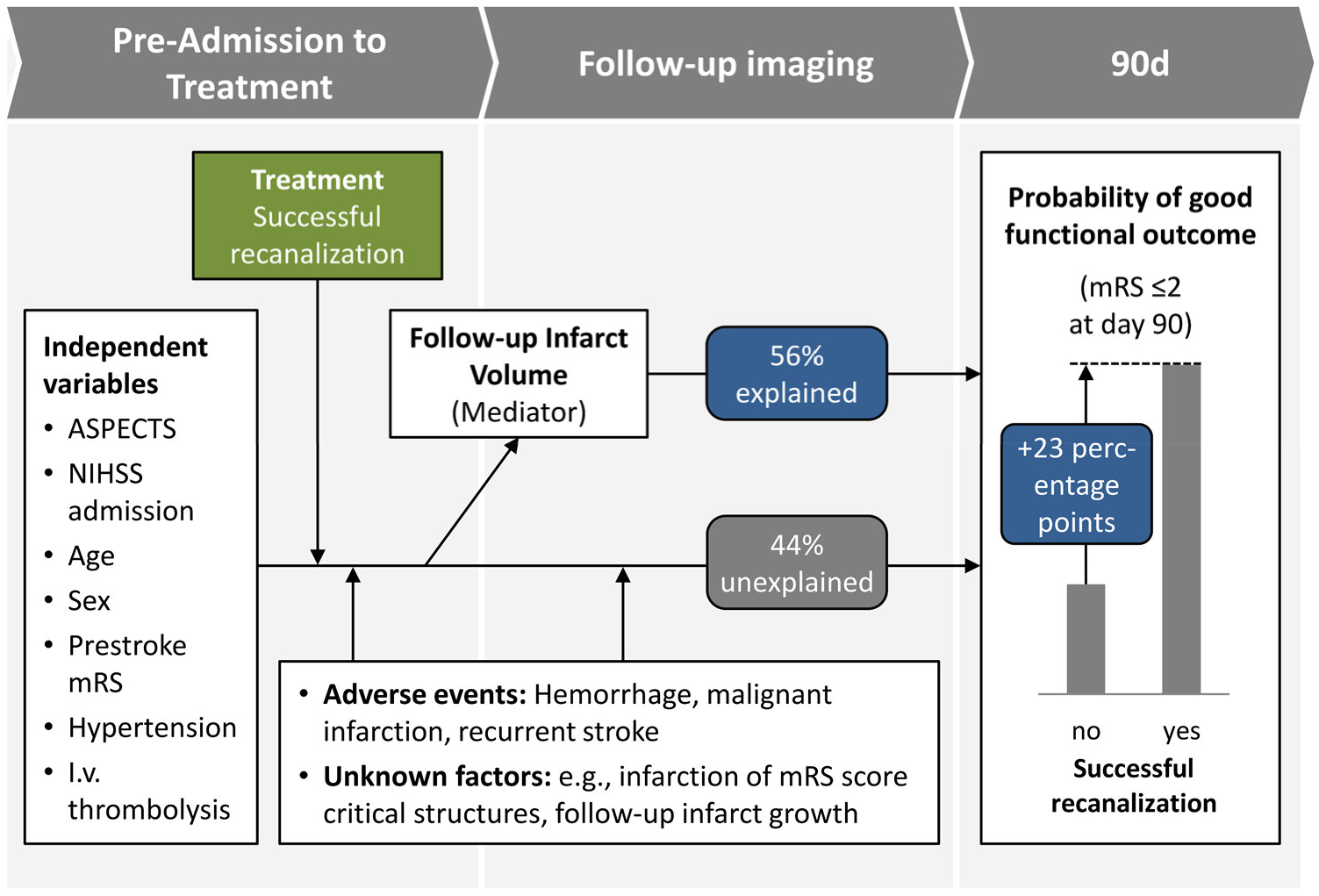
Mediation model layout and result overview. Abbreviations: mRS: Modified Rankin Score; SD: Standard Deviation; NIHSS: National Institutes of Health Stroke Scale; ASPECTS: Alberta Stroke Program Early CT Score; Tici: Thrombolysis in Cerebral Infarction.

All requirements for mediation relationship according to Baron and Kenny ^14^ were fulfilled: Regression analysis of the direct path confirmed significant association of the independent predictors and the mediator FIV with the probability for good outcome. Furthermore, significant coefficients were observed for the regression of the mediator on the independent predictors. All effect metrics were significant at p-value < 0.001. The layout of the mediation model is depicted in Figure 2.

Results of the mediation analysis suggest a 23 percentage points (pp) increase of probability of good functional outcome (95% confidence interval (CI): 16pp – 29pp) in patients with successful recanalization. 13pp (95% CI: 9pp – 18pp) of the effect was mediated by FIV. 56% (95% CI: 38% - 78%) of the treatment effect was explained FIV reduction following successful recanalization with MT (Table 2).

**Table 2:** Mediation analysis results.

| Outcome: 90d mRS ≤ 2 | | | |
| --- | --- | --- | --- |
| Treatment: TICI ≥ 2b vs. $<$ 2b | | | |
| Mediator: Follow-up infarct volume (FIV) |  |  |  |
| Increase of probability of 90d mRS ≤ 2 | Estimate | 95% CI | p-value |
| Total effect | 23.1pp | [16pp; 29pp] | $<0.001$ |
| Direct effect | 10.2pp | [4pp; 17pp] | $<0.001$ |
| Effect mediated by FIV | 13.0pp | [9pp; 18pp] | $<0.001$ |
| % of total effect mediated by FIV | 55.9% | [38%; 78%] | $<0.001$ |

Abbreviations: mRS: Modified Rankin Score; TICI: Thrombolysis in Cerebral Infarction; FIV: Follow-up infarct volume

## DISCUSSION

Our analysis based on 429 patients with acute ischemic stroke confirms that FIV after MT is associated with functional outcome and partially mediates the relationship between MT and functional outcome at 90 days after successful recanalization. Our results indicate that 56 % of the total treatment effect of successful recanalization in patients with anterior circulation stroke is explained by FIV reduction, suggesting that additional characteristics and mechanisms such as infarct location, extent and severity of brain tissue damage, delayed neurological improvement capacity, effective rehabilitation, prevention of late adverse events and other unknown factors account for the remaining 44% of the treatment effect.

Several studies have previously addressed the association of FIV with functional outcome after an occlusion of the anterior circulation. In concordance with our results, it was demonstrated that FIV after MT in anterior circulation strokes serves as a reliable predictor for 90 day functional outcome ^4–7^. Those studies also found that patients treated with MT had significantly smaller FIVs compared with controls treated with best medical treatment.

Furthermore, four recent studies examined if the relationship between treatment and functional outcome was mediated by FIV ^2,8–10^. Two of these studies ^2,9^ reported that FIV did not mediate the relationship between treatment and functional outcome. In the other two studies ^8,10^, only a small proportion (12-14 %) of the treatment effect was reported to be explained by FIV reduction. In contrast, our analysis indicates that FIV reduction explains 56% of the improvement in functional outcome after successful recanalization. However, these studies evaluated the effect of MT versus best medical treatment on functional outcome and did not assess the effect of successful recanalization. In comparison, our study only included patients treated with MT and evaluates the effect of successful recanalization on FIV, long term functional outcome and the extend of mediation through FIV. Based on the observed higher percentage of explained treatment effect, it could be concluded that in comparison to MT vs best medical treatment, the effect of successful recanalization might be more clearly and pathophysiologically more directly associated with FIV reduction.

It is noteworthy that in our study the beneficial effect of successful recanalization on functional outcome was mainly explained by FIV reduction. It therefore can be assumed that FIV reduction is a significant factor leading to improved outcome in patients undergoing MT. However, our study also holds important information on the remaining mismatch between radiological and clinical outcome measures after treatment as 44% of the improvement in outcome is not explained by FIV reduction. For example, FIV cannot capture external effects not related to MT, such as efficiency of rehabilitation, adverse events due to comorbidities or respiratory infections, which are known as significant determinants of higher morbidity and mortality in stroke patients ^20^. Moreover, FIV does not consider the exact location and the eloquence of the affected brain areas. Also, former studies have reported significant infarct growth between 24 hours and 1 week of follow-up ^21^ and little is known about infarct progression after the 1-week follow-up period. These prolonged pathophysiological processes might reduce the predictive power of FIV for long term functional outcome.

Our results corroborate established pathophysiological assumptions and confirm the value of infarct volume as imaging endpoint in clinical trials.

Observed improvement of functional outcome of 23pp after successful recanalization corresponding to a number-needed to treat of 4.3 reflects numbers reported for large RCTs investigating the effect of MT ^22–28^.

Our study has limitations. First, selecting the last scan for each patient for FIV assessment could have led to a bias, as patients with adverse events post treatment might have had higher chances of receiving late imaging. Second, the analysis is based on a binarized functional outcome measure, which might oversimplify the problem. However, binarization of 90d mRS with regards to good functional outcome (mRS ≤ 2) is an established approach for the definition of clinical endpoints. In addition, patients with a history of stroke might present with relatively poor clinical outcomes, weakening the overall association of FIV with functional outcome. However, pre-mRS was included as covariate in pour model. Third, to generate valid results from mediation analysis, unmeasured confounding must not exist between parameters in the hypothetical causal model. This is a strong assumption, especially when considering that the many interconnected biological processes are not fully understood and might vary on patient-specific level. However, we can expect that this unmeasured confounding effect is reduced, as relevant predictors of functional outcome after MT were considered during the design of the causal model. Fourth, the proportions of the association of successful recanalization with functional outcome explained by FIV could never reach the theoretical value of 0% or 100% due to the underlying assumptions of the hypothetical causal model.

## Conclusions

56% of the improvement of functional outcome after successful recanalization is explained by FIV reduction. Results corroborate established pathophysiological assumptions and confirm the value of infarct volume as imaging endpoint in clinical trials. 44% of the improvement in outcome is not explained by FIV reduction and reflects the remaining mismatch between radiological and clinical outcome measures.

## Data Availability

The data that support the findings of this study are available from the GSR-ET registry and the corresponding author, restrictions may apply to the availability of these data.

## ACKNOWLEDMENTS

Authors acknowledge the German Stroke Registry (GSR) investigators and the GSR steering committee: Prof. Dr. med. Joachim Röther (Asklepios Klinik Altona, Hamburg); Prof. Dr. med. Bernd Eckert (Asklepios Klinik Altona, Hamburg); Dr. med. Michael Braun (Bezirkskrankenhaus Günzburg); Prof. Dr. med. Gerhard F. Hamann (Bezirkskrankenhaus Günzburg); PD Dr. med. Eberhard Siebert (Charité –Benjamin Franklin und Campus Charité); Prof. Dr. med. Christian Nolte (Charité –Benjamin Franklin und Campus Charité); Dr. med. Sarah Zweynert (Charité - Campus Virchow Klinikum, Berlin); Dr. med. Georg Bohner (Charité - Campus Virchow Klinikum, Berlin); Prof. Dr. med. Jörg Berrouschot (Klinikum Altenburger Land); Dr. med. Albrecht Bormann (Klinikum Altenburger Land); Dr. med. Christoffer Kraemer (Klinikum Lüneburg); PD Dr. med. Hannes Leischner (Klinikum Lüneburg); Dr. med. Jörg Hattingen (KRH Klinikum Nordstadt Hannover); Dr. med. Martina Petersen (Klinikum Osnabrück); Prof. Dr. med. Florian Stögbauer (Klinikum Osnabrück); PD Dr. med. Boeckh-Behrens (Klinikum r.d.Isar); Dr. med. Silke Wunderlich (Klinikum r.d.Isar); Dr. med. Alexander Ludolph (Sana Klinikum Offenbach); Dr. med. Karl-Heinz Henn (Sana Klinikum Offenbach); Prof. Dr. med. Christian Gerloff (UKE Hamburg-Eppendorf); Prof. Dr. med. Jens Fiehler (UKE Hamburg-Eppendorf); Prof. Dr. med. Götz Thomalla (UKE Hamburg-Eppendorf); Asklepios Klinik Altona, Hamburg (UKE Hamburg-Eppendorf); Dr. med. Anna Alegiani (Asklepios Klinik Altona, Hamburg); Dr. med. Maximilian Schell (UKE Hamburg-Eppendorf); PD Dr. med. Arno Reich (Uniklinik RWTH Aachen); Prof. Dr. med. Omid Nikoubashman (Uniklinik RWTH Aachen); Prof. Dr. med. Franziska Dorn (Uniklinik Bonn); Prof. Dr. med. Gabor Petzold (Uniklinik Bonn); Prof. Dr. med. Jan Liman (Klinikum Nürnberg); Dr. med. Jan Hendrik Schäfer (Uniklinik Frankfurt/ Main); Dr. med. Fee Keil (Uniklinik Frankfurt/ Main); Prof. Dr. med. Klaus Gröschel (Universitätsmedizin Mainz); Dr. med. Timo Uphaus (Universitätsmedizin Mainz); Prof. Dr. med. Peter Schellinger (Universitätsklinik Johannes Wesling Klinikum Minden); Prof. Dr. Jan Borggrefe (Universitätsklinik Johannes Wesling Klinikum Minden); Dr. med. Steffen Tiedt (Uniklinik München (LMU)); PD Dr. med. Lars Kellert (Uniklinik München (LMU)); PD Dr. med. Christoph Trumm (Uniklinik München (LMU)); Prof. Dr. med. Ulrike Ernemann (Universitätsklinik Tübingen); PD Dr. med. Sven Poli (Universitätsklinik Tübingen); Prof. Dr. med. Christian Riedel (Universitätsmedizin Göttingen); PD Dr. med. Marielle Sophie Ernst (Universit ätsmedizin Göttingen).

